# Recommendations for an Optimal Model of integrated case detection, referral, and confirmation of Neglected Tropical Diseases: A case study in Bong County, Liberia

**DOI:** 10.1101/2022.02.01.22269709

**Authors:** Tiawanlyn G. Godwin-Akpan, Shahreen Chowdhury, Emerson J. Rogers, Karsor K. Kollie, Fasseneh Z. Zaizay, Anna Wickenden, Georgina V.K. Zawolo, Colleen B.M.C. Parker, Laura Dean

**Affiliations:** AIM Initiative/American Leprosy Mission, Greenville, South Carolina, USA; Department of International Public Health, Liverpool School of Tropical Medicine, Liverpool, United Kingdom; Neglected Tropical Disease Program, Ministry of Health, Oldest Congo Town, Monrovia, Liberia; Neglected Tropical Disease Program, Ministry of Health, Oldest Congo Town, Monrovia, Liberia. Current Address: REDRESS, Oldest Congo Town, Monrovia, Liberia; AIM Initiative/American Leprosy Mission, Greenville, South Caroline, USA. Current Address: Effect Hope, Markham, ON, Canada; Countdown Liberia, Pacific Institute for Research and Evaluation, Capitol Hill, University of Liberia, Monrovia, Liberia; Research Unit, Ministry of Health, Liberia, Oldest Congo Town, Monrovia, Liberia

## Abstract

**Background:** People affected by Neglected Tropical Diseases (NTDs), specifically leprosy, Buruli ulcer (BU), yaws, and lymphatic filariasis, experience significant delays in accessing health services, often leading to catastrophic physical, psychosocial, and economic consequences. Global health actors have recognized that Sustainable Development Goal 3:3 is only achievable through an integrated inter and intra-sectoral response. This study evaluated existing case detection and referral approaches in Liberia, utilizing the findings to develop and test an Optimal Model for integrated community-based case detection, referral, and confirmation. Finally, this study evaluates the efficacy of implementing the Optimal Model in improving the early diagnosis of NTDs.

**Methodology/Principal Findings:** The study used mixed methods, including key informant interviews, focus group discussions, participant observation, quantitative analysis, and reflexive sessions to evaluate the implementation of an Optimal Model developed through this study. The quantitative results from the testing of the optimal model are of limited utility. The annual number of cases detected increased in the twelve months of implementation in 2020 compared to 2019 (pre-intervention) but will require observation over a more extended period to be of significance. Qualitative data revealed essential factors that impact the effectiveness of integrated case detection. Data emphasized the gendered dynamics in communities that shape the case identification process, such as men and women preferring to see health workers of the same gender. Furthermore, the qualitative data showed an increase in knowledge of the transmission, signs, symptoms, and management options amongst CHW, which enabled them to dispel misconceptions and stigma associated with NTDs.

**Conclusion/Significance:** This study demonstrates the opportunity for greater integration in training, case detection, rereferral, and confirmations. However, the effectiveness of this approach depends on a high level of collaboration, joint planning, and implementation embedded within existing health systems infrastructure. Together, these approaches improve access to health services for NTDs.

**Author Summary:** Global health professionals and stakeholders have advocated for integration across diseases and sectors to improve the success of public health interventions. This advocacy has also impacted NTDs programs globally. NTDs interventions are becoming more integrated than disease-specific activities to maximize limited resources, improve coverage and access to healthcare services. However, documentation on the effectiveness of integrated approaches to improve access to healthcare services is minimal. This study evaluated existing case detection and referral approaches in Liberia, utilizing the findings to develop and test an Optimal Model for integrated community-based case detection, referral, and confirmation. Finally, this study evaluates the efficacy of implementing the Optimal Model in improving the early diagnosis of NTDs. The results provide evidence of the benefits of an integrated approach and the programmatic challenges to achieve the goal of improving access to health services for persons affected by NTDs.

## Introduction

Globally, people affected by Neglected Tropical Diseases (NTDs), specifically leprosy, Buruli ulcer (BU), yaws, and lymphatic filariasis, experience significant delays in receiving diagnosis and treatment; this is due to multiple factors, including stigma, a lack of knowledge, accessibility, and gender (1). However, compounding these challenges is the lack of the availability of appropriate disease management and disability inclusion (DMDI) services of reasonable quality in the community available through the health system (1,2). Social stigmatization and discrimination are very high for people with NTDs due to the visible physical impairments caused by many NTDs. Therefore, remaining hidden in their communities is a typical response to stigma (3,4). This stigma and lack of awareness of NTDs further contribute to the poor health-seeking behavior of people with NTDs (2,5–7). The time between symptoms, diagnosis, and treatment determines the treatment regimen and illness outcome (8). Early diagnosis and management reduce the risk of significant physical, economic, and psychosocial impacts of NTDs (9). Therefore, addressing the barriers to early case detection is essential to attaining universal health coverage and reducing the disease burden caused by NTDs; this includes a shift from a passive approach to case detection to an active model. The World Health Organization (WHO) has emphasized active case finding as part of the five essential intervention packages to prevent, control, and eliminate NTDs (10,11). Integrating active case finding into national NTD programs will promote active surveillance instead of passive data collection to ensure no one is left behind.

NTDs affect more than a billion people worldwide, impacting low-income populations in Africa, Asia, and the Americas (8). NTDs are a diverse group of 20 tropical diseases broken into two major groups: preventive chemotherapy and transmission control (PCT) and innovative and intensified disease management (IDM). PCT NTDs are managed at the community level by mass drug administration (MDA). At the same time, IDM NTDs require early diagnosis and management on an individual basis (12). NTDs are an equity tracer for the Sustainable Development Goals (SDGs), with specific targets. SDG sub-goal 3:3 states, “to end the epidemics of AIDS, tuberculosis, malaria, and neglected tropical diseases and combat hepatitis, water-borne diseases, and other communicable diseases by 2030” (13). Global health actors have recognized that this is only achievable through an integrated inter and intra sectoral response (14).

This article’s integration of NTDs refers to the joint planning, implementation, and evaluation of activities across programs to achieve common goals (15). Following the launch of the first WHO Roadmap for NTDs (8), emphasis and prioritization have been placed on providing preventative Mass Drug Administration (MDA) for PCT NTDs. There is far less emphasis and attention on diagnostic, curative, and morbidity management services for IDM NTDs (excluding leprosy). The (PCT) NTDs have been integrated since the early 2010s through MDA with evidence of success. However, the (IDM) NTDs lag as they require considerable resources, including personnel with skills and financial support (9,16,17).

Consequently, with restricted funding to specific IDM disease conditions (e.g., Buruli ulcer or leprosy), IDM NTD interventions have been vertical and cannot reach affected persons (10). As a result, the global commitment to “leave no one behind” cannot be achieved without support to NTD programs to identify, diagnose, and manage IDM NTDs(18). Furthermore, delay in diagnosis, treatment, and referral can lead to the ongoing transmission of those NTDs of epidemic concern, disability and morbidity challenges, and increased economic hardship for underprivileged families.

Liberia was one of the first countries to adopt an integrated approach to the case management of (IDM) NTDs to improve access and universal health coverage for people with NTDs. This approach responded to lessons learned from the Ebola Virus Disease (EVD) outbreak and two national policies: “Essential Package of Health Services” and “Investment Plan for building a resilient health system,” which were developed to build resilient health system (19,20). In line with these policies, the NTDs and the National Tuberculosis and leprosy control programs, along with several national and international partners, developed a “National Strategic Plan for the Integration of Case Management of NTDs (CM-NTDs) in Liberia (2016–2020)”(21). For Liberia context, CM-NTDs are synonymous with (IDM) NTDs. The plan extends the National NTDs Masterplan, which follows the WHO NTDs Masterplan template for planning PCT and IDM NTDs interventions. The strategic plan for integrated case management aligns with the four strategic priorities of the WHO NTDs Masterplan. These priorities are: 1) government ownership, advocacy, coordination, and partnerships, 2) resource mobilization and planning for results, 3) access to interventions and treatment improved and system capacity building, and 4) monitoring, surveillance, and operational research enhanced (21). The plan includes strategies to integrate BU, leprosy, clinical manifestations of lymphatic filariasis (hydrocele and lymphedema), and yaws for case detection, referral, confirmation, and management control to eliminate NTDs by 2020 (21,22). The fifteen counties in Liberia are all co-endemic for the targeted diseases requiring case management (21). Due to funding restraints in 2017, the NTD program launched the integrated plan for case management in five counties: Bong, Bomi, Lofa, Maryland, and Nimba, alongside the integration of national-level activities including budgeting, performance monitoring, data management, and drug supply chain. The NTD Program implemented multiple and diverse community-based case detection models in the pilot counties to increase early case detection, as shown in Fig 1. However, the effectiveness of these differing models in increasing patient access to care and early diagnosis had not been documented.

**Fig 1.**
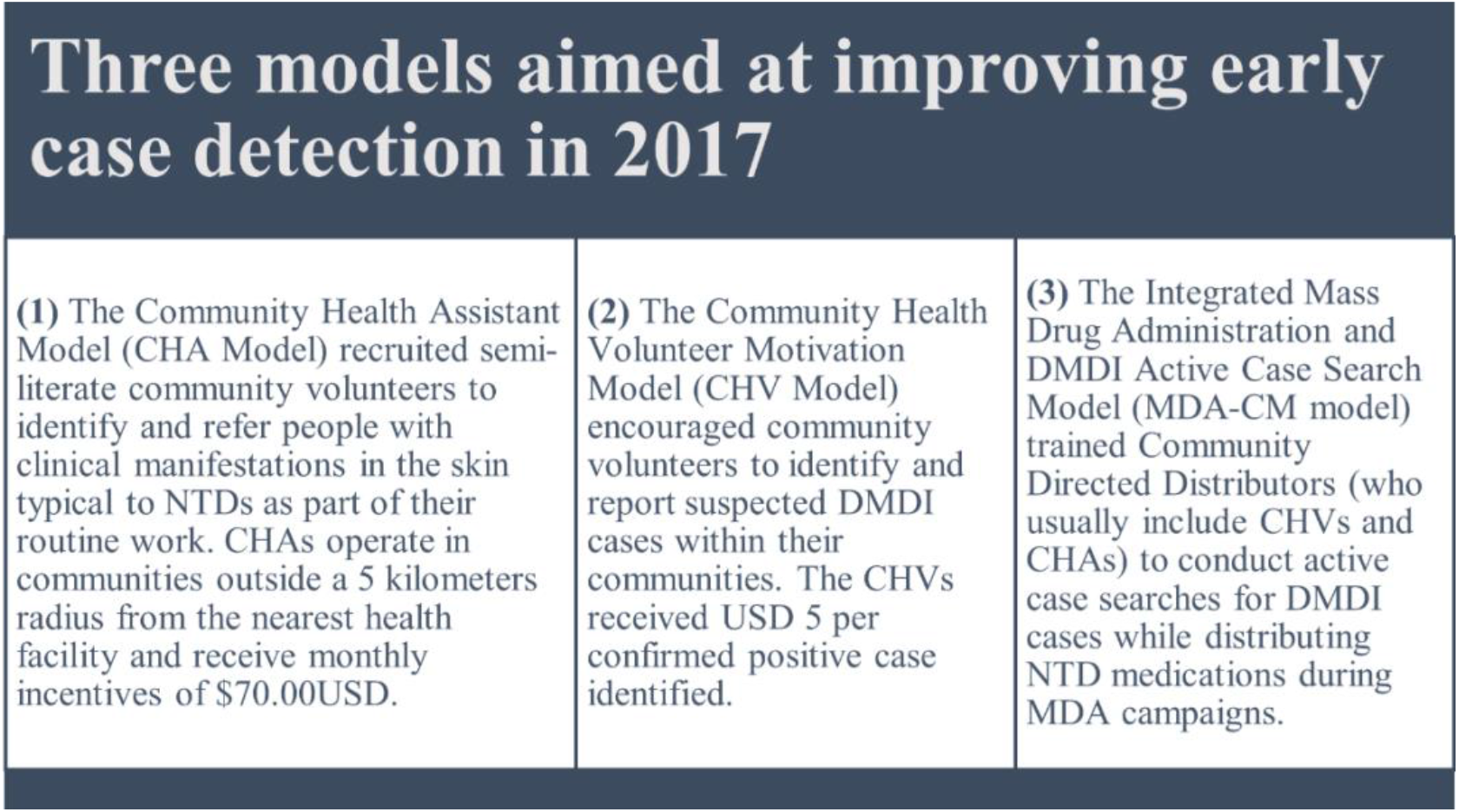
NTDs program community-based case detection models implemented through the integrated case management approach in the five pilot counties in 2017.

This study aimed to evaluate existing approaches to case detection and referral in Liberia using an action research cycle, use the findings to develop an optimal model for integrated community-based case detection, referral, and confirmation. Finally, this study evaluates the efficacy of the optimal model in improving early diagnosis NTDs. This paper presents findings from the formative research phase, describes the optimal model produced, and the key learnings from its implementation over twelve months.

### The Optimal Model

The Optimal Model was developed in partnership with the National Ministry of Health NTD team, the Bong County Health Team, people affected by NTDs, and community health workers with support from national NTD program partners (American Leprosy Missions, Liverpool School of Tropical Medicine, University of Liberia Pacific Institute for Research and Evaluation) through COR-NTD funding. There was a collaboration between the national program, lower levels of the health system, and persons affected by NTDs during the entire development process. The model was developed in two phases over nine months: phase one, the formative phase, and phase two, the planning phase. During phase one, we conducted a participatory qualitative evaluation of the program’s three models of case detection to identify strengths and weaknesses. We used the results to design an Optimal Model for case detection. This phase drew on qualitative participatory methods, including key informant interviews and focus group discussions with health workers and affected persons on understanding their experiences of existing case detection approaches. During phase two, the planning phase, a two-day workshop was held in Monrovia, Liberia, with stakeholders including persons affected by NTDs, community health workers (CHWs), Bong County Health team, NTD program, and other health stakeholders from the central, county, and district levels to collaboratively develop the optimal model based on the findings from phase one. The findings included vital themes such as inconsistencies and delays in remuneration, geographical barriers, opportunity costs throughout pocket expenditure of CHWs on transport and calls, and insufficient training and supervision of DMDI services. Challenges regarding gender, disability, and other axes of social disadvantage patients face were also explored to understand how they affect case detection; poverty, social isolation, stigma, and gendered access were highlighted as critical issues. The optimal model included comprehensive training on integrated approaches to identify, refer, diagnose and manage NTDs at the community level. The training included a referral process, supervision structure, and incentive packages. The Optimal Model developed is summarized in Fig 2 and detailed in S1 File.

**Fig 2.**
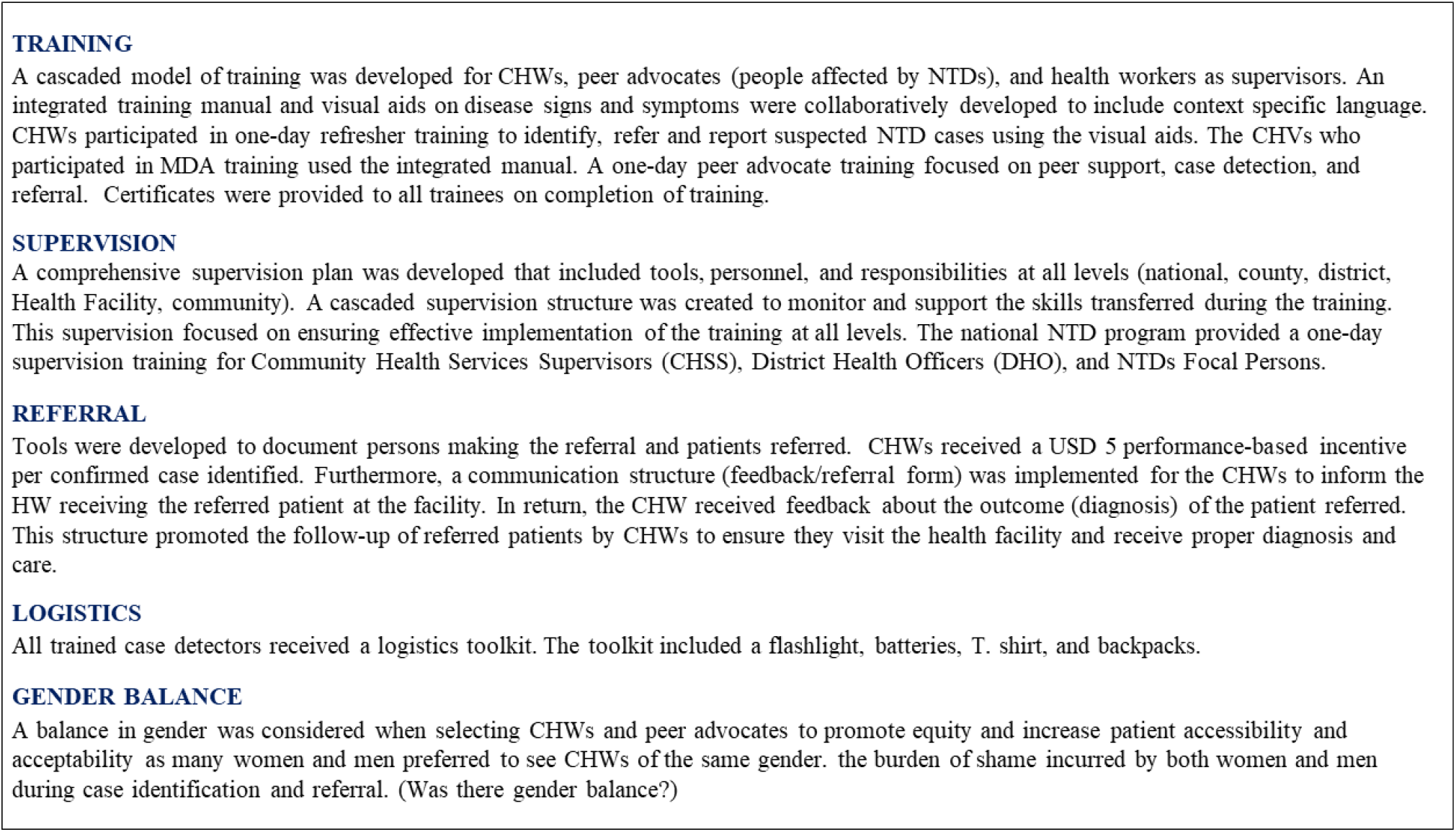
The optimal model for case detection, referral, and confirmation of NTDs

## Materials and Methods

### Ethics Statement

This assessment was approved by the University of Liberia-Pacific Institute for Research & Evaluation (UL-PIRE) Institutional Review Board (Protocol number: NTDSC 167D) (S2 File).

### Consent and Data Collection Procedure

Informed consent was obtained from each participant in the assessment before FGDs, KIIs, and reflexive sessions. The participants had an opportunity to ask questions about the study and the implications of their participation. Participants’ confidentiality was assured, and the right to withdraw from the research at any time of the interview. Upon completing the consent process, each participant received a copy of their signed consent form (S3 File). Anonymizing names and other identifiers maintained confidentiality.

### Study Setting

Bong County was purposively selected as the study setting for this research based on the following criteria: Bong County is one of the five counties of the integrated CM-NTDs pilot; co-endemic for all priority diseases; has mixed success in terms of case reporting across its nine health districts (i.e., some have identified more cases than others); and all three models of case detection (Fig 1) have been implemented in this setting. Furthermore, Bong shares borders with two integrated CM-NTDs implementation counties (Nimba and Lofa). Patients may travel between these counties for treatment (23). The study was conducted in Bong County, Liberia (Fig 3).

**Fig 3.**
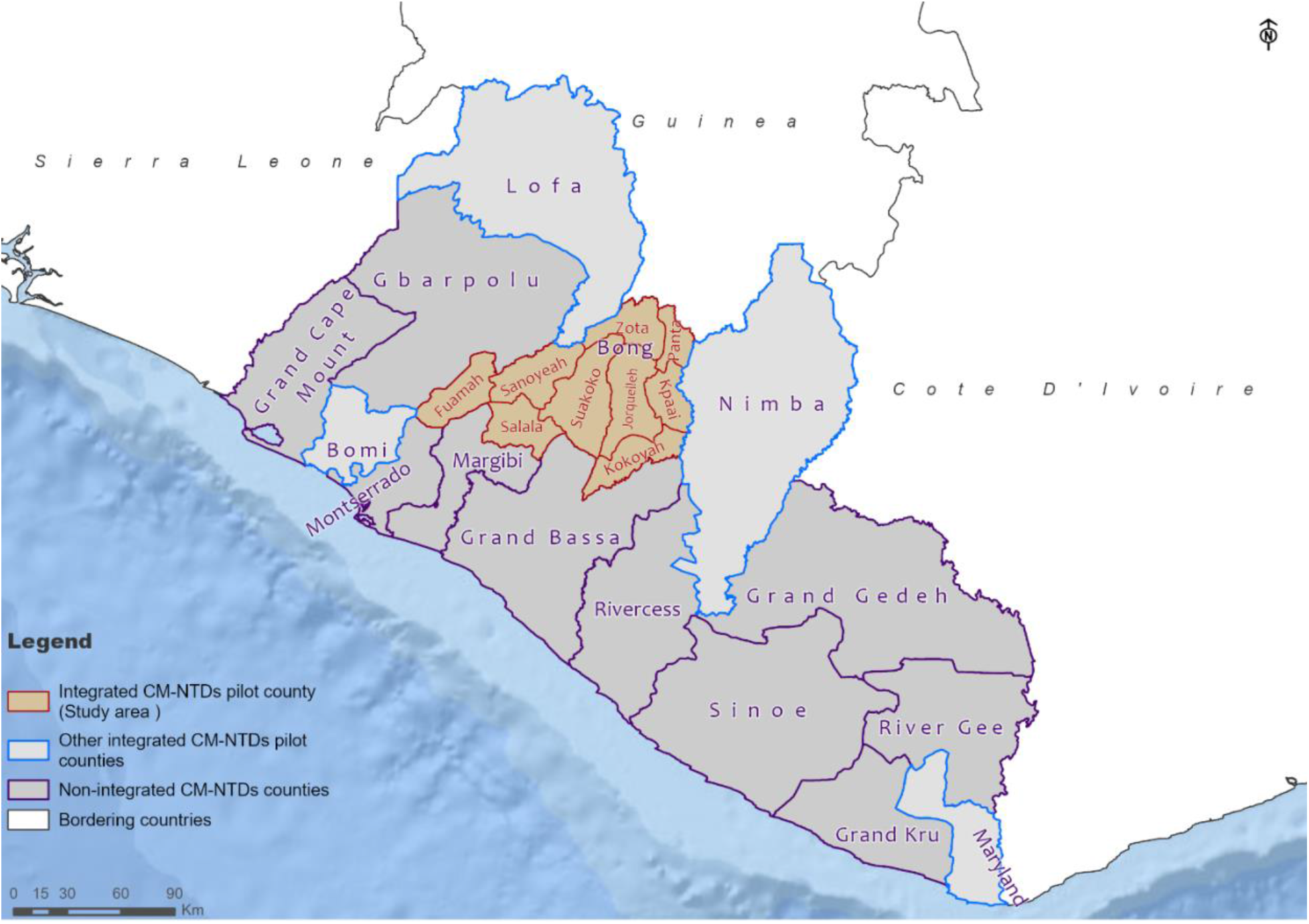
Map of Liberia showing the study site, Bong County, and other integrated CM-NTDs pilot counties

### Study Design

The study uses a Community-Based Participatory Research (CBPR) approach comprising phases depicted in Fig 4. The CBPR approach promotes the equitable participation of community members, researchers, and all stakeholders throughout the research process (24). During phases 1-2 (nine months period), the optimal model was developed (as described above) and implemented during phase 3 (one-year period). The findings presented in this study were identified in phase four (four months period), the observation and reflection phase, based on the activities in phases 1,2, and 3. Phase four used a mixed-methods approach to evaluate the implementation of the optimal model, which is both a process and outcome evaluation.

**Fig 4.**
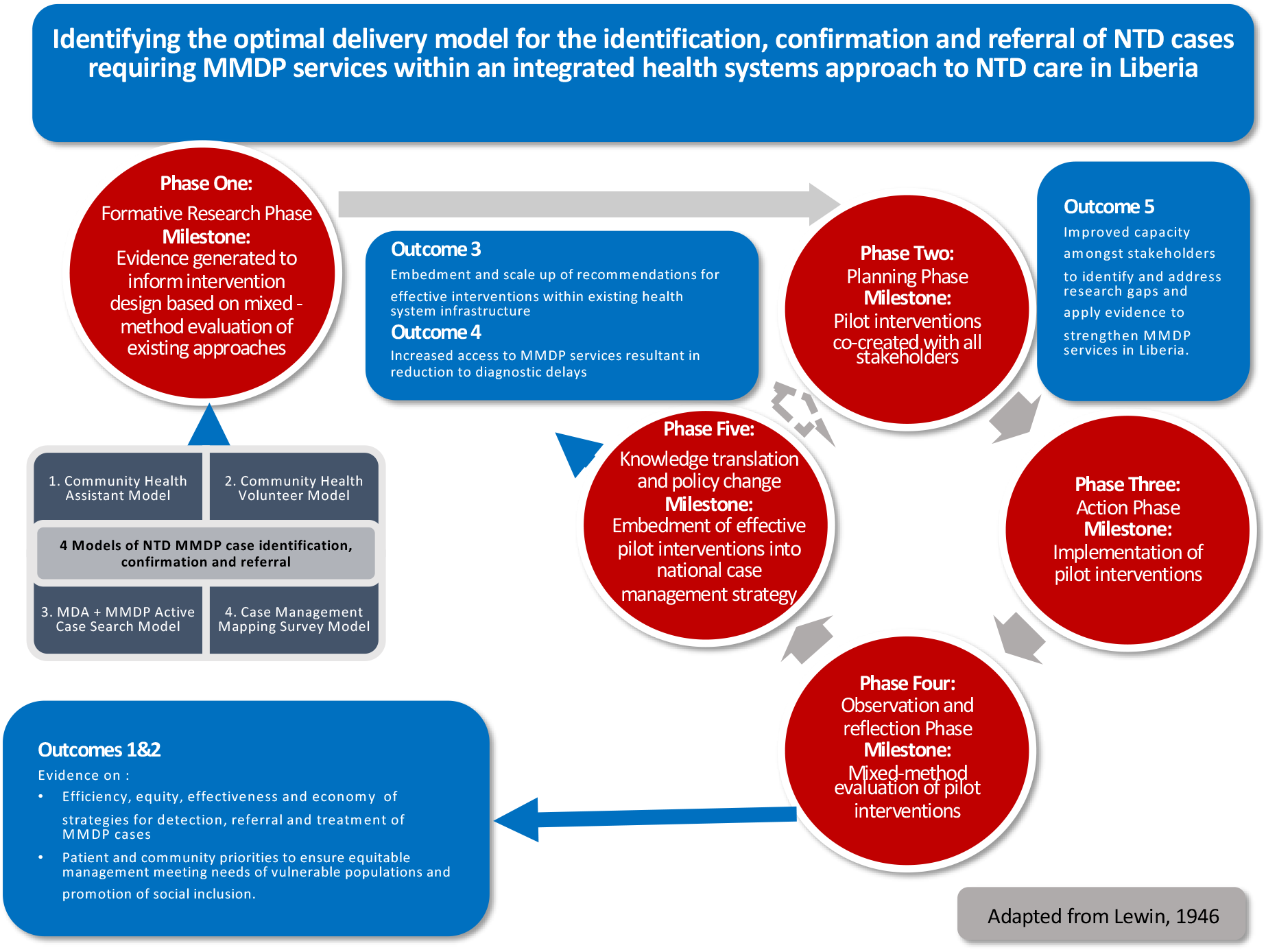
Conceptual Framing, Theory of Change and Community Based Participatory Research Phases

### Qualitative Methods

During phase four, we conducted a mixed-method evaluation which included key informant interviews, focus group discussions, participant observation, reflexive sessions. Participants were purposively selected based on their role within delivering the optimal model to affected populations and for maximum variation in gender and age. Furthermore, participants were selected from all health system levels and had experience in integrated case detection, referral, and confirmation of NTDs or benefited from NTD services.

### Data Collection

Table 1 presents the different data collection methods for the evaluation and participants.

**Table 1.** Overview of Data Collection

| Data Collection | Number of Sessions | Number of Participants | Total Participants |
| --- | --- | --- | --- |
| *Process Evaluation* |  |  |  |
| A reflexive session with the Research team | 1 | 5 | 5 |
| A reflexive session with the County Health team | 1 | 2 | 2 |
| Participant observation at CHW training | 1 | 760 | 1 |
| Participant observation at supervision training | 1 | 48 | 1 |
| Participant observation at patient advocate training | 1 | 36 | 1 |
| *Outcome Evaluation* |  |  |  |
| Key Informant Interviews with CHSS | 4 | 1 | 4 |
| FGD with CHWs at health facilities | 4 | 5 | 20 |
| FGD with patient advocates after training | 1 | 5 | 5 |
| Quantitative Analysis of Health Systems Data | N/A | N/A | N/A |
| **Total** | **14** | **21** | **39** |

### Key Informant Interviews

Four health facilities in Bong were purposively selected based on the geographical variation. A CHSS trained to supervise CHW in the community in each facility was interviewed as a key informant, using an interview guide. We were particularly interested in their experiences and perspectives of training and supervision, specifically, how the optimal model has impacted case identification, referral, and confirmation at different health system levels.

### Focus Group Discussions

Five FGDs were conducted with community health workers and peer advocates based on their roles and training. We were interested in their training experience and how cases were identified, referred to, and confirmed using the optimal model at different health system levels. Separate group discussions were held with CHWs and patient advocates. Each focus group discussion had between 5-8 participants.

### Participant Observation

Participant observation was conducted at the training of CHWs, supervisors, and peer advocates. A structured observation grid was used to document reflections and observations of interactions, participation, and training content.

### Reflexive Sessions

Reflexive sessions involving a participatory group discussion to reflect on the optimal model’s design, delivery, and evaluation were conducted with the core research team and the County Health Team. The core research team remotely participated in a reflexive session over Skype. In contrast, a reflexive session with the county health team was conducted at the health facility, including the NTD focal person and the Community Health focal person in Bong County. The reflexive session included key themes on collaboration, participation and power, the use of evidence, and reflections on the challenges, strengths, and sustainability of the optimal model.

### Data Analysis

All interviews were digitally recorded and transcribed in English. All data were anonymized and stored securely in a password-protected database. Debriefing sessions with data collectors focused on key findings, identifying saturation themes, and refining inquiry lines. Data were analyzed thematically using a framework approach which involved the research team reading and re-reading the transcribed data. A coding framework was developed, transcripts were coded using NVivo software, with emerging themes discussed and coding refined. The coded transcripts were charted and summarized in narratives for each theme.

### Quantitative Methods

During the optimal model pilot test period (January-December 2020), pre-existing and routine NTDs program data reporting tools were used to collect quantitative data on the utilization of NTDs services. Existing health systems data was disaggregated by gender and other socio-demographic factors and utilized to collate information about the impact of the optimal model. Descriptive comparisons are made between the optimal model intervention period (2020) and the preceding year (2019) against key SMART performance-based evaluation indicators. These indicators include; the number of confirmed cases for NTD morbidity (number of confirmed hydrocele cases, number of confirmed lymphedema cases, percentage of BU cases identified with categories 1 and 2 lesions, percentage of newly identified Leprosy patients presenting with grades two disabilities), and number of confirmed CM-NTD cases per disease.

### Data Analysis

The research team compiled a list of NTD patients diagnosed over the twelve months recorded in the health facility registers in an excel spreadsheet. The individual line list of patients was analyzed using SPSS version 24.

## Results

### A. QUALITATIVE DATA: Optimal model design and delivery

#### Inclusion and collaboration

Partnership and collaboration promoted ownership of the process of developing, implementing, and evaluating the optimal model as stakeholders were involved in all stages. The research and implementation team all agreed that collaboration with the County teams, community health workers, and persons affected by NTDs strengthened the design of the optimal model in order to ensure the language was context-specific and easily understandable at the community level.

> *’We could bring everyone on board, and everyone gave their collective views on how the intervention phase should be implemented, which gave birth to the development of training materials, development of IEC materials, how to motivate the community and health workers…search for cases.’ (National Program Staff, Reflexive Session)*

#### Training

Key strengths of the training included pictorial guides and accessible language, which aided case detection at the community level. The inclusion of stigma education helped CHWs improve disease understandings and dispel misconceptions and stigma associated with NTDs. The training also increased knowledge of NTD transmission, signs and symptoms, and management options. The additional training provided has expanded the program’s reach as health facilities that were previously dormant diagnosed NTDs cases in 2020.

> *“Yes, from the training, I began to realize that if you have a patient with NTDs, people will begin to stigmatize them; oh, this person got elephantiasis I don’t want to get some. But from the training, my thoughts began to tell me that that person is important no matter of their conditions, so I should learn to live with the person just as how I have to live as human beings” (CHW, FGD, Bong)*
>
> *’During the development of the revised training modules, it was really nice to have the community of workers make inputs…the language really changed… the community health workers gave a lot of, consideration of all of the diseases in that ‘you have to say it in this way, and that really made the revised modules really rich’ (Researcher, Reflexive Session)*

The length of training was a key challenge. One day for training was too short, and many CHWs requested longer training sessions or refresher training. Within this pilot, the duration of the COR-NTD training deviated from the original optimal model, which was initially planned to last for two days due to budget constraints. As a result, participants reported feeling rushed and needed more time on the referral forms to practice filling them out correctly.

> *“The training is for…it’s not supposed to be for twenty-four hours. It is supposed to be for forty-eight hours or seventy-two hours because the notes’ training is so bulky. You can’t use one day to put everything in your brain. You have to take some time to study. Even though they had pretest and posttest. It was okay but it has to take some days to get your brain reminding you of it.” (CSS, KII)*

#### Trust and Rapport with Communities: The essential role of peer advocates

Peer advocates are affected by NTDs who have received DMDI services and played a unique role in case-finding activities. Many peer advocates expressed how sharing their experiences has increased trust and encouraged more people to attend the health facility. Peer advocates reported relating well to other persons affected to address stigma and encourage them to seek treatment.

> *’My experience is I talk with the person, and I tell him I too have suffered from these kinds of diseases and the Phebe hospital took care of it easily, and he should not be scared…and I too have gone under such an operation, and nothing happened….The good thing I experienced about the training was to talk with those that got the sickness like me, and they got treated and got well like me because I have been in the hospital myself before.’ (Peer Advocates, FGD, Bong)*

#### Gender Balance

Gendered dynamics shaped the case identification process in communities. Men and women felt more comfortable discussing their conditions and being screened by CHWs of the same gender. However, the training sensitized case detectors to consider gender preferences by involving relatives or friends during case detection; many CHWs described that this resulted in affected persons being more likely to cooperate.

> *’Though the person will find it very difficult to express their condition to you as an opposite gender because traditionally they do not feel secure.*
>
> *’In identifying cases with women, it’s kind of a little quite different. If she’s married and perhaps she has a problem, if you must be successful in handling that problem, you must firstly engage the husband making sure to let him know what the problem the woman has so he can know what is your role too in the community. So, in my experience on the field, before engaging somebody wife, if she has a problem, I must also let the husband know that I came on the field and this is my job. This is what your wife has and this is what I want to do. He must approve before I can go on with my function.’ (CHWs, FGD, Bong)*

#### Supervision

The supervision cascade was effective, as CHWs and CHAs reported feeling supported and encouraged due to the feedback received from supervisors. Some forms were not filled out correctly in the first supervision round, but this improved with the second round. According to many CHSS, supervisions were conducted daily in different communities where community awareness meetings were held. Coordination amongst the different levels of health cadres in the supervision cascade was a key implementation strength. This coordinated approach through the use of supervision forms at all levels and a detailed supervision plan was not included in the previous models.

> *‘Those who can come visit us we can really enjoy them because some of the thing them there I can’t understand it. So when he comes he can teach me and it can go well with me. For example, I just use to put the date I will not put nothing on it. But my supervisor when she comes she can say no that’s not how you can do it. Do it so. When even you do not find the case, you are supposed to do it so. So it can really encourage me. It building me up to do the work now.’ (CHW, FGD, Bong)*

However, logistical challenges such as limitations in transportation, gasoline, and communication cards were mentioned by supervisors as limitations to optimal supervision. Financial constraints in paying for drugs and lack of drugs in facilities were crucial challenges. However, this was highlighted by NTD Program staff as being a national issue and not reflective of the optimal model.

> *‘Our CHSS does not have support like gasoline for his bike and he most often used his own money to purchase gasoline to go on the field to check on us. I think the government needs to find ways to assist him because that’s the only way he too will be able to supervise us.’ (CHW, FGD, Bong)*

#### Referral

Improvements in referral and reporting as a result of the optimal model were demonstrated through the referral structure. Tools such as referral forms, counter referral forms, and the NTDs ledger improved reporting. The referral process entailed CHWs and peer advocates filling in referral forms in the community and referring the patient to the health facility for confirmation and treatment. When confirmed positive, the patients were put on treatment, with a counter referral form filled by the health facility and sent back to the community as feedback to the CHW who referred the case.

> *‘But for now, things being better for the past year. Because cases that I have being getting in recent time, as soon I call the county focal person if that case specimen supposed to be collected I get in touch. He will try to connect and try to collect the specimen. As soon that case is confirmed, the drugs will immediately come. As we even speak I get patient in the facility the facility that are on their treatment and they are improving.’ (CHSS, KII, Bong)*

However, some CHSS and CHWs described challenges in the referral of patients, which included limitations of forms, financial constraints, and lack of drugs at the health facilities.

#### Remuneration

The performance-based incentive package was motivating, as mentioned by many CHWs although some CHWs experienced delays in receiving the USD 5 and incentive packages. Certificates based on the number of cases referred were also considered a source of encouragement. Other sources of motivation include pride in their roles, success stories of patients receiving treatment, and recognition in the community.

> *’Anytime we find case for me I brought cases here I got bag. I got five dollars. So those are motivating us in order for us to move further. I was motivated really. Because if that after the training, my first case that I found I received bag, five dollars with flashlight.’ (CHW, FGD, Bong)*
>
> *’What we think could work better is that because most of the time, this five-dollar issue can’t really become a problem in the facility. When the CHV report the cases, the focal person will call you and say the case is confirmed; you will give your feedback to the CHV in the community. They expect their five-dollar at that time but at time it will take long time before the five dollar. All those can make them feel reluctant with the work’ (CHSS, KII, Bong)*

### B. Quantitative Data: Optimal model delivery

Across all diseases, the annual number of cases diagnosed based on clinical manifestation has increased compared to the previous year (Fig 5). This was particularly noticeable for hydrocele and suspected Buruli ulcer cases, with more than twice the number of cases reported in 2020 compared to 2019 (Fig 6). Cases reported in 2020 are from across all health facilities within Bong County. This is the first year that geographic coverage appears to have reached all health facilities since the original rollout of integrated case detection in 2016/17. Cases were detected at an earlier stage of disease progression. A significant proportion of Buruli ulcer cases have been identified with grade I and grade II lesions in 2020 compared to 2019 (Fig 7). Grade 1 lesions would indicate greater capacity and earlier diagnosis in Buruli ulcers. The proportion previously was almost entirely grade II lesions. However, results related to Buruli ulcer should be interpreted with caution due to the limitations in PCR confirmation due to the COVID-19 pandemic and reduced supplies to Liberia.

**Fig 5.**
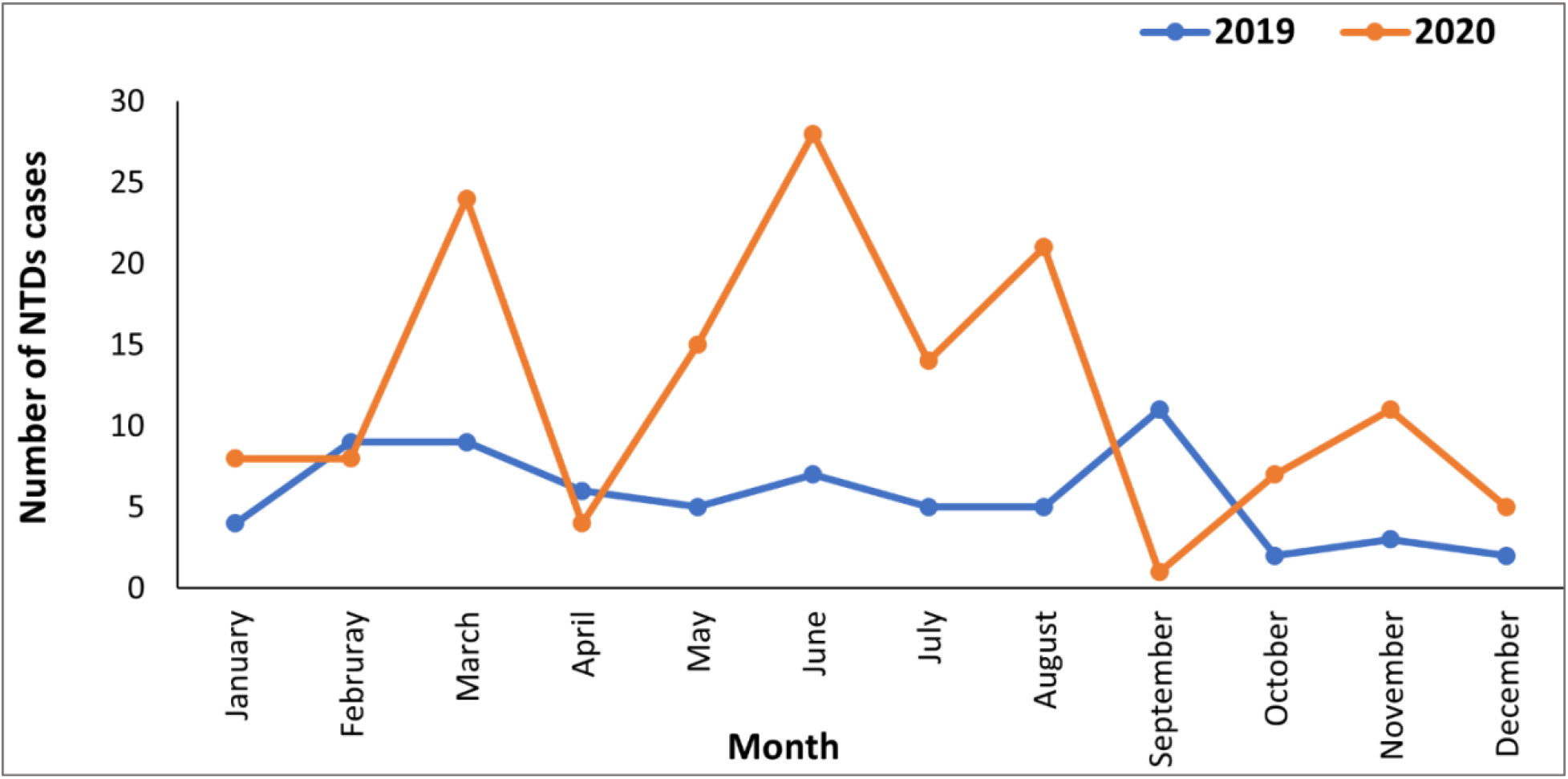
Comparison in cases reported in 2019 (year proceeding the optimal model) and 2020 (year of optimal model implementation)

**Fig 6.**
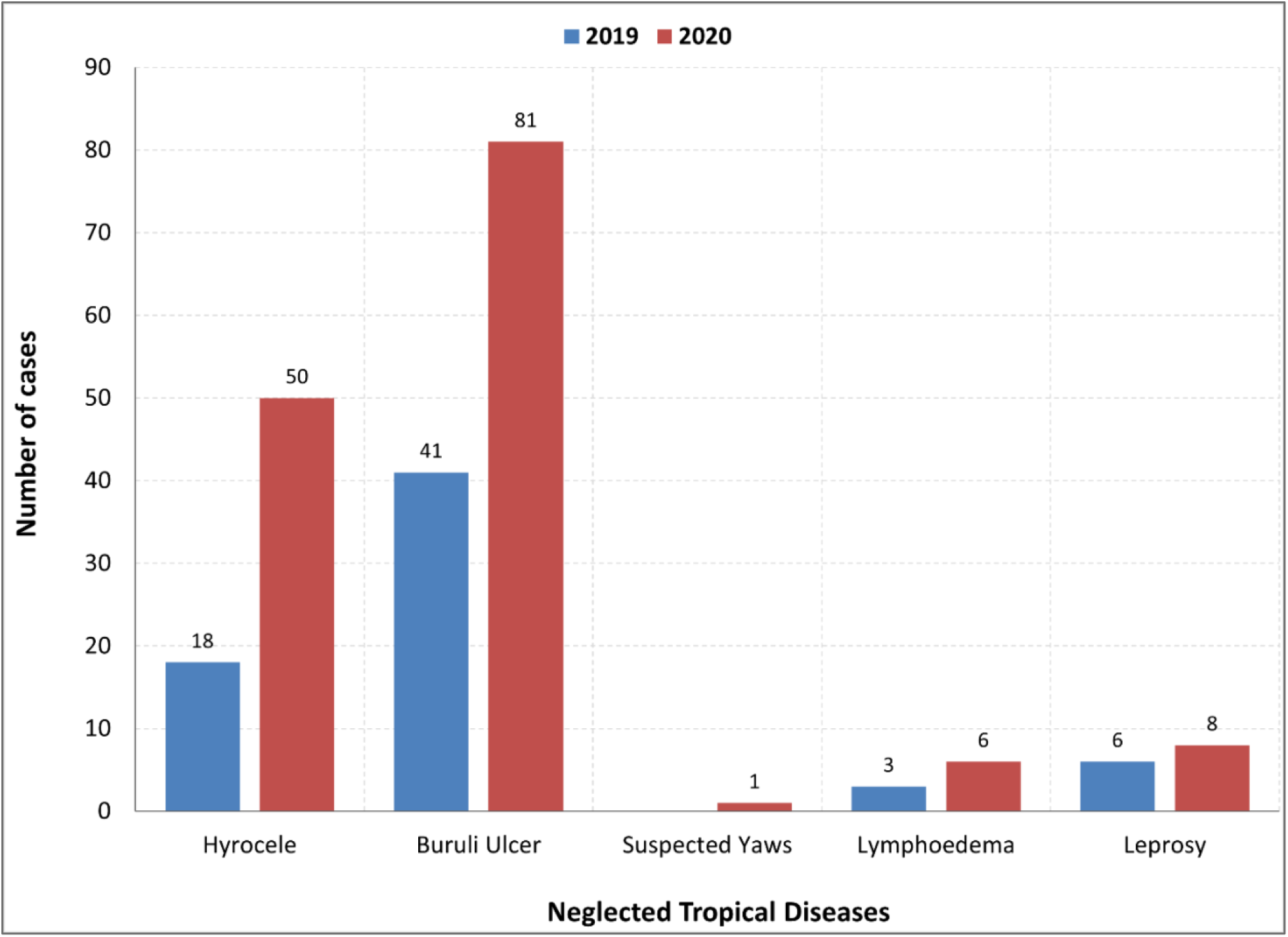
Annual comparison of the number of CM NTDs Cases per disease

**Fig 7.**
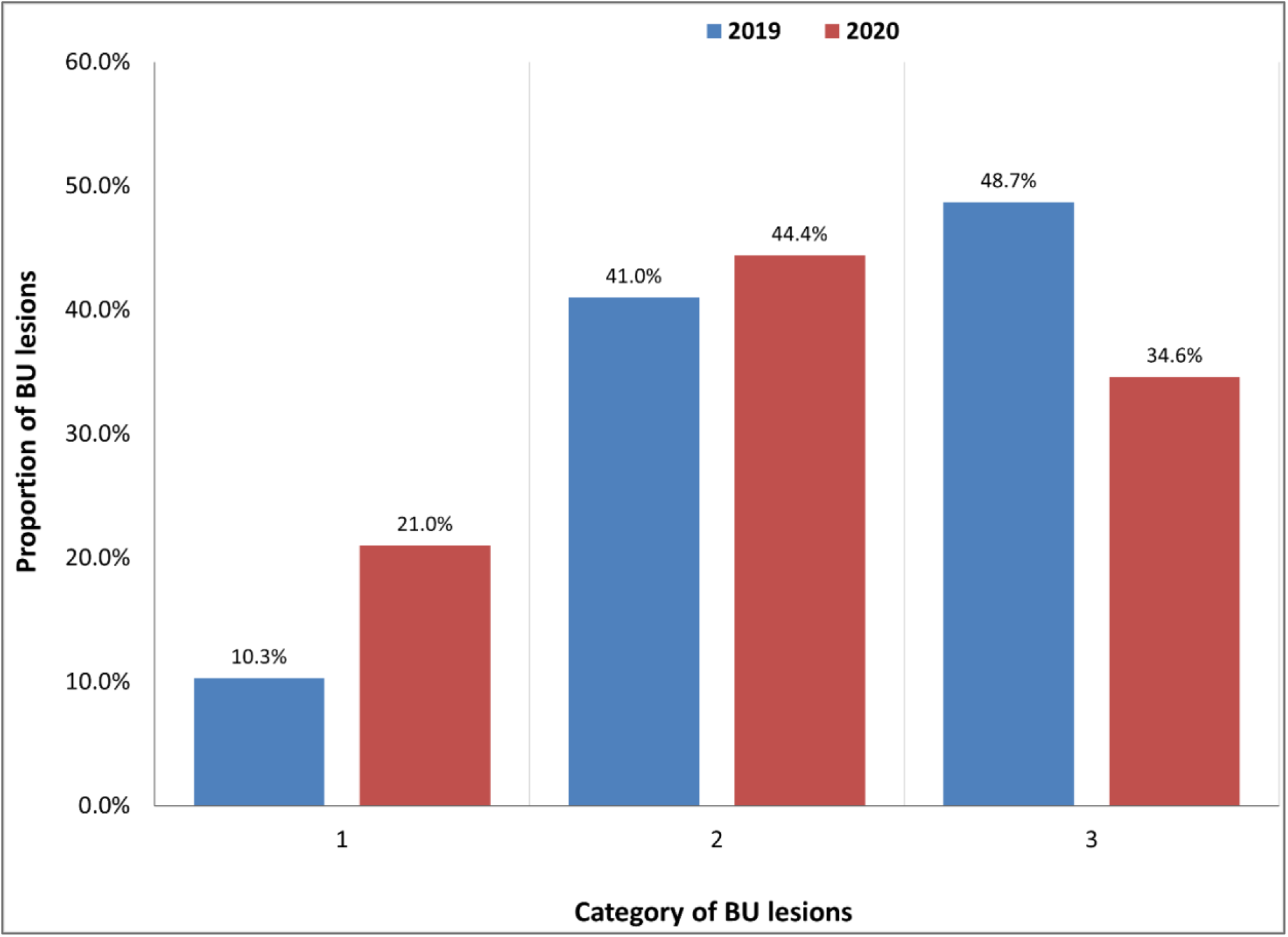
Percentage of newly diagnosed BU patients per year with category I, II, or III lesions at point of detection

The Optimal Model also shows an increase in the number of health facilities throughout the county that reported cases (Fig 8). However, many patients reported some level of movement limitations at diagnosis due to ulceration.

**Fig 8.**
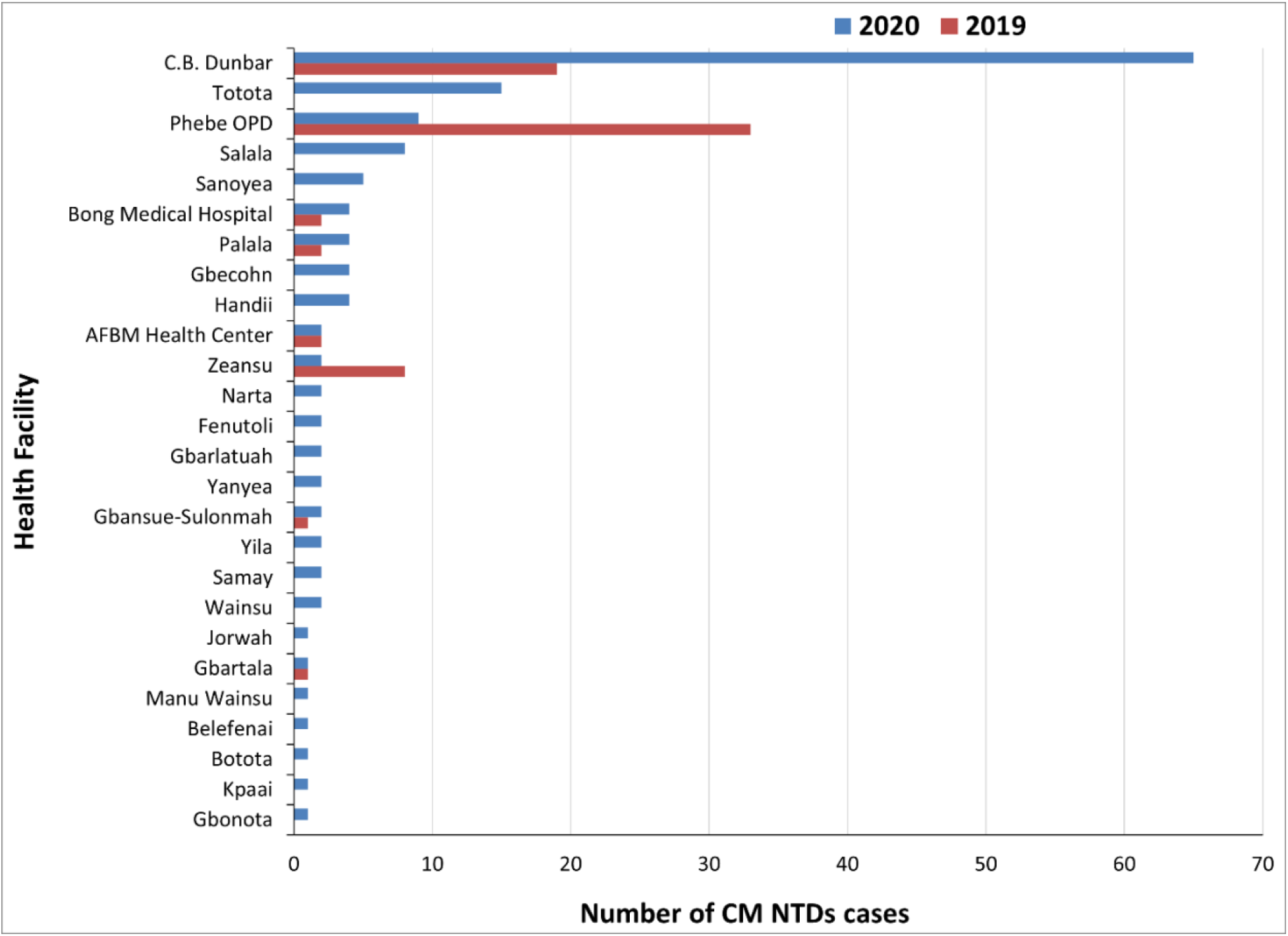
Comparison of the Annual number of CM-NTDs Cases detected per Health Facility in Bong County.

By 2020, more health facilities reported CM NTDs cases referred by CHVs who have been trained and supported through the Optimal Model package than in 2019. Although the number of cases reported in 2019 and 2020 per health facility was deficient, several health facilities reported CM-NTDs cases for the first time in 2020 due to the Optimal Model (Fig 8).

Impact of gender on equitable access – a higher proportion of male cases than females was identified across diseases (Fig 9). Five hundred ninety-five males and 109 females participated in the case detection and referral training from the nine health districts in Bong County (Table 2). The CHW training included both males and females between the ages of 25-60 from catchment communities of all the health facilities in Bong County. However, significantly more men than women were in this training, as reflected in the cases detected (Fig 9). A more significant proportion of cases identified as males is shared across all disease conditions across 2019 and 2020, excluding leprosy, where case detection appears equal (Fig 9).

**Fig 9.**
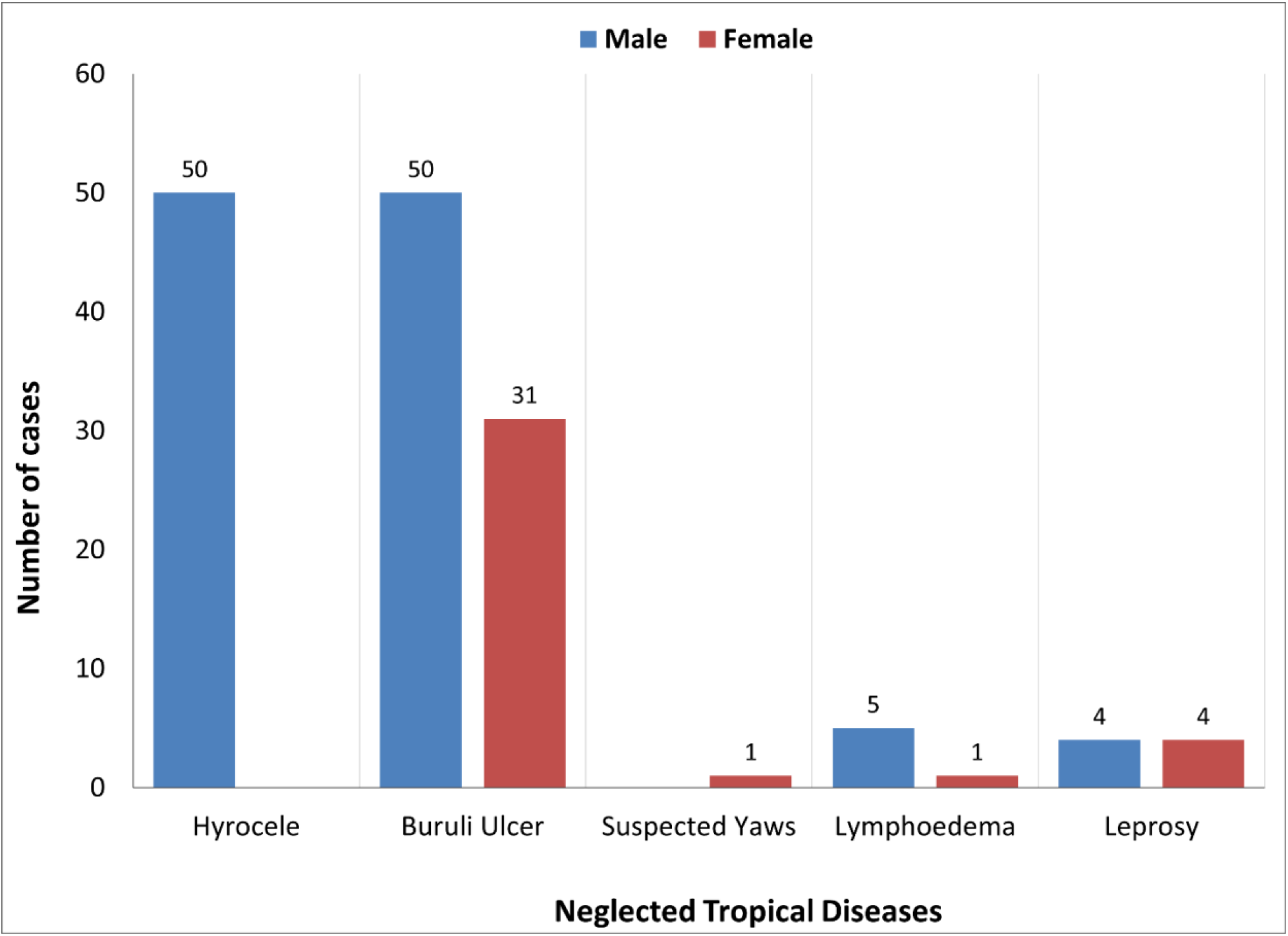
Case Management of NTDs cases disaggregated by sex per disease condition in 2020, Bong County, Liberia

**Table 2.** Composition of participants trained, disaggregated by role and gender

| Role of Trainee | Male | Female | Total |
| --- | --- | --- | --- |
| CHWs | 595 | 109 | 704 |
| CSS | 29 | 28 | 57 |
| Peer Advocates | 35 | 15 | 50 |
| **Total** | **659** | **152** | **811** |

## Discussion and Conclusion

One of the main objectives of the Optimal Model was to support the earlier identification of NTDs requiring case management to reduce morbidity and disability. For Buruli ulcer, this was successful, with a significant proportion of cases being identified with grade I and grade II lesions in 2020 compared to 2019. This result emphasizes that cases are detected at an earlier stage of disease progression. The training improved case finding across all diseases as the annual number of cases diagnosed had increased in 2020 compared to 2019 in Bong County.

This study, to our knowledge, is the first to focus on identifying an optimal model for integrated case detection, referral, and confirmation of NTDs in Liberia. Several studies have been conducted on the integrated case management of NTDs strategy in Liberia, focusing on patient perception, the outcome of care, and case detection (22,25,26). There have been findings on the effectiveness of active case search in increasing the number of cases compared to reliance on passive surveillance at the health facility for specific diseases (27,28). However, the findings from this study support improved access to care for affected persons through early case detection.

Integrating different disease conditions within a single training reduced barriers to care for persons affected by NTDs. The collaboration between different cadres of health workers to develop a comprehensive training manual supported enhanced and early case detection for NTDs affected persons with poor health-seeking behavior. Our study found that integrating these disease conditions into one manual increased CHWs’ knowledge to conduct community awareness to reduce stigmatization of persons affected by NTDs. The observations in this study are similar to Mitja *et al.*, who found that “the success of an integrated approach will rely on well-trained health workers and village volunteers being able to correctly identify multiple skin conditions” within one interaction (11). The CHWs benefited from the collaboration across diseases and amongst the different levels of the health system. The knowledge gained and confidence in their ability to provide valuable information on NTDs to community members and identify suspect cases was due to the participation of various stakeholders in the development of the training tools and the design of the optimal model.

Alongside integrated training, ensuring the right people receive this training and are well motivated is essential. For example, our study found that Buruli ulcer and hydrocele cases reported in 2020 were twice reported in 2019. CHWs reported that this was significantly due to the integrated training and the frequent supervision and incentives, which motivated them to continue active case search and community awareness. The on-spot mentoring and follow-up during supervision by CHSS provided an opportunity for CHWs to gain more knowledge and confidence to identify cases. This result supports the findings of studies conducted in the Republic of Benin, Cameroon, and Nigeria who also found an increase in the number of cases reported with early signs of the disease through referral by trained community health workers (29–32). In addition, our study prioritized the inclusion of peer advocates as case detectors to build trust with community members and motivate affected persons to visit the health facility for proper diagnosis. Youtsu also found that integrated surveillance requires the training of local health workers (16). Peer advocates’ personal experience with the disease conditions and their understanding of the local community helps reduce stigma and encourage positive healthcare-seeking behaviors. Program decision-makers need to carefully select who delivers the activities to impact program outcomes.

Gendered access to patients to identify and refer was a key challenge emphasized by qualitative and quantitative data. The gender disparity in the number of cases diagnosed was linked to the sex of those who received the training. CHWs reported seeking permission from spouses or family members to screen suspected cases of the opposite sex due to cultural norms and fear of stigmatization. This finding is consistent with findings from India that women delayed hospital visits awaiting permission from their husbands or guardians (33). In order to address gender inequities in CM-NTDs interventions, programs must invest in frontline health workers to improve communication techniques to reduce shame and stigma as barriers to healthcare seeking (34). Despite the commitment to gender equity within the research, it was impossible to achieve gender parity in training as the community health workforce in Liberia is predominantly male. National programs must proactively advocate for gender balance in the recruitment of CHWs to ensure that those conducting activities truly represent the gender makeup of such a community. Additionally, integrating gender in all stages of planning, tool development, testing, implementation, and evaluation will improve the equity and impact of case finding.

### Limitations

The delivery of this study was limited by the impact of COVID-19 due to partial lockdowns and social distancing restrictions. Liberia confirmed its first COVID-19 case on March 16, 2020. On April 8, 2020, the President declared a 21-day renewable national state of emergency for residents of all fifteen counties until further notice. This meant no inter-county travels between the capital and other counties and a 3 p.m. to 6 a.m. curfew rather than a total lockdown. During this time, residents were permitted to leave their homes only to procure food or health items, an activity limited to one person per household for a maximum of one hour within his or her local area. This state of emergency was extended two weeks until July 21, 2020. Therefore, case detection activities were suspended, resulting in the low number of cases reported in the quantitative results. The fluctuation in the number of cases detected between February to September 2020 corresponds with the increase in Covid-19 cases in Bong County. Case detection began in February 2020, with high numbers reported after completion of the integrated training in January; however, there were significantly fewer cases reported with the first lockdown between March and April. In contrast, NTDs cases reported between August to September 2020 also decreased when no new COVID-19 was reported in Bong county. This is due to the increased workload for CHWs as other health programs began to resume activities after the decrease in COVID-19 cases and the end of lockdown measures.

Many CHWs mentioned how COVID-19 impacted their work in case identification and referrals. People feared attending health facilities for fears of contracting COVID-19. This resulted in reduced regular community visits, reduced referrals, limited health talks. To mitigate the impact of COVID-19 on the case detection activities, infection prevention and control measures were implemented. Hand sanitizers and face masks were provided to CHWs to reduce their fear of interacting with community members during case detection when possible.

Furthermore, many CHWs were recruited and trained to conduct COVID-19 surveillance, which was done simultaneously with NTDs case detection and awareness activities. COVID-19 also affected the work of peer advocates. While peer advocates’ significant role in case finding was recognized, the duration of their case-finding activities was far less than that of CHWs. As a result, the impact of their work cannot be quantitatively measured for case-finding activities.

### Recommendations for Health System Strengthening

Implementation of the optimal model did come with challenges, which are to be anticipated when integrating interventions into emerging health systems. Despite the global impetus towards integration, restrictions on donor funding limited the scope of some activities, e.g., length of training and supervision., greater flexibility concerning the scope and integration within the training of multi diseases would enable more effective case finding for NTDs. Similarly, a significant limitation was the logistical support required to implement a robust supervision model. The importance of supportive supervision was repeatedly emphasized; however, the investment in this area was insufficient. Based on these challenges as documented in the results, it is recommended that the time allocated for training should be increased. For example, a three-day training was suggested to allow more time to practice the referral and reporting forms. Supervisors need logistical support to provide on-spot mentoring for CHWs on the supervision forms. Provision of adequate copies of the referral forms to health facilities is suggested to improve documentation and reporting of the referral process. The remuneration system of cash, certificates, and other rewards should be strengthened to avoid delays and encourage CHWs in their work.

The challenges documented in the study also highlight the financial constraints and barriers faced by NTDs programs in implementing such an integrated model that requires skilled human resources and logistics for routine activities. Funding allocation will significantly impact the sustainability and scaling up of the optimal model. Implementing the optimal model is largely dependent on donor support; components of the optimal model, such as the incentive package, will be harder to sustain without financial support. With the documented success of integrating active case finding and collaboration at all health system levels through this study, national budgets should prioritize funding for integrated approaches to the case management of NTDs. Low reliance on donor funding will enable the NTDs program to make independent decisions and lead in the prioritization of critical interventions.

### Investment in integration

This study has shown how integrated case detection of NTDs is pivotal to achieving the global target to leave no one behind(18). The second WHO NTDs roadmap 2021-2030 prioritizes integrated approaches to achieving new global targets(36). The increase in the number of cases detected and diagnosed at an early stage of the disease through the optimal model outlined in this study enables evidence-based advocacy with policymakers. Optimal models for case detection, referral, and confirmation of suspect NTD cases rely on integrated approaches to training, supervision, referral, and remuneration that are embedded within existing health systems infrastructure. Equity and inclusion are also critical, particularly the gender of health workers and the inclusion of affected persons. The global NTD target SDG 3:3 can only be achieved through optimal case detection, referral, and confirmation models. Together, these approaches improve access to health services, thus reducing morbidity associated with NTDs.

## Data Availability

All data produced in the present work are contained in the manuscript

## Acknowledgment

We would like to extend our gratitude to the following people, without whose support this project would not have been successful. Many thanks to the Ministry of Health Liberia, the data collection team comprised of Mohammad Dunbar and Otis Kpadeh, and field drivers for their commitment and endurance during the data collection process. Many thanks to the Ministry of Health NTD program monitoring and evaluation team and the UL-PIRE for the approval to conduct the research, and finally, our most tremendous thanks to the national stakeholders, clinical staff, CHAs, CHVs, CDDs, patients, and communities for their time to participate in this study. Finally, we appreciate Paul Saunderson and Stefanie Weiland for their review of earlier drafts of the manuscript, which contributed immensely to the final draft.

## Supporting Information

S1 File. Detailed Optimal Model for case detection, referral, and confirmation

S2 File. Ethical approval

S3 File. An example of consent form administered

S1 Appendix. Supervision Tools

S2 Appendix. Certificate of Training

S3 Appendix. Training Manual

S4 Appendix. Referral forms

## Notes

### Competing Interest Statement

The authors have declared no competing interest.

### Funding Statement

This study was funded by the Department for International Development (DFID) through the Task Force for Global Health-COR-NTD; however, the funders had no role in study design, data collection and analysis, decision to publish, or preparation of the manuscript.

### Author Declarations

University of Liberia-Pacific Institute for Research and Evaluation Institutional Review Board (UL-PIRE IRB)

## References

1. Engels D. Neglected tropical diseases in the Sustainable Development Goals. Vol. 387, The Lancet. Lancet Publishing Group; 2016. p. 223–4.

2. Ramchandra Atre S, Girish Rangan S, Prabhakar Shetty V, Gaikwad N, Furdoon Mistry N. Perceptions, health-seeking behavior and access to diagnosis and treatment initiation among previously undetected leprosy cases in rural Maharashtra, India. Leprosy review. 2011;82(3):222.

3. Prochazka M, Timothy J, Pullan R, Kollie K, Rogers E, Wright A, et al. “Buruli ulcer and leprosy, they are intertwined”: Patient experiences of integrated case management of skin neglected tropical diseases in Liberia. PLoS Neglected Tropical Diseases. 2020 February 5;14(2).

4. Hofstraat K, van Brakel WH. Social stigma towards neglected tropical diseases: a systematic review. International Health. 2016 Mar 3;8(suppl 1):53–70.

5. Nicholls PG, Chhina N, Bro AK, Barkataki P, Kumar R, Withington SG, et al. Factors contributing to delay in diagnosis and start of treatment of leprosy: analysis of help-seeking narratives in northern Bangladesh and in West Bengal, India. Leprosy Review. 2005;76(1):35–47.

6. Weiss MG. Stigma and the social burden of neglected tropical diseases. PLoS Neglected Tropical Diseases. 2008 May 14;2(5):237.

7. Van GY, Onasanya A, van Engelen J, Oladepo O, Diehl JC. Improving access to diagnostics for schistosomiasis case management in Oyo state, Nigeria: Barriers and opportunities. Diagnostics. 2020 May 1;10(5):328.

8. World Health Organization. ACCELERATING WORK TO OVERCOME THE GLOBAL IMPACT OF NEGLECTED TROPICAL DISEASES: A ROADMAP FOR IMPLEMENTATION. 2012.

9. Mieras LF, Anand S, van Brakel WH, Hamilton HC, Martin Kollmann KH, Mackenzie C, et al. Neglected Tropical Diseases, Cross-Cutting Issues Workshop, 4–6 February 2015, Utrecht, the Netherlands: meeting report. International Health. 2016 March 3;8(suppl 1):7–11.

10. Mitjà O, Marks M, Bertran L, Kollie K, Argaw D, Fahal AH, et al. Integrated Control and Management of Neglected Tropical Skin Diseases. PLOS Neglected Tropical Diseases. 2017 January 19;11(1).

11. Ortu G, Williams O. Neglected tropical diseases: exploring long term practical approaches to achieve sustainable disease elimination and beyond. Infectious Diseases of Poverty. 2017 Dec 27;6(1):1–2.

12. Rosenberg M, Utzinger J, Addiss DG. Preventive Chemotherapy Versus Innovative and Intensified Disease Management in Neglected Tropical Diseases: A Distinction Whose Shelf Life Has Expired. PLOS Neglected Tropical Diseases. 2016 April 14;10(4).

13. TRANSFORMING OUR WORLD: THE 2030 AGENDA FOR SUSTAINABLE DEVELOPMENT. New York, NY; 2015.

14. World Health Organization. Department of Control of Neglected Tropical Diseases. Integrating neglected tropical diseases into global health and development : fourth WHO report on neglected tropical diseases. Geneva; 2017.

15. Boisson S, Engels D, Gordon BA, Medlicott KO, Neira MP, Montresor A, et al. Water, sanitation and hygiene for accelerating and sustaining progress on neglected tropical diseases: a new Global Strategy 2015–20. International Health. 2016 March 3;8(suppl 1).

16. Yotsu R. Integrated Management of Skin NTDs—Lessons Learned from Existing Practice and Field Research. Tropical Medicine and Infectious Disease. 2018 Nov 14;3(4).

17. Simpson H, Quao B, van der Grinten E, Saunderson P, Ampadu E, Kwakye-Maclean C, et al. Routine Surveillance Data as a Resource for Planning Integration of NTD Case Management. Leprosy Review. 2018;89(3):178–96.

18. Fitzpatrick C, Engels D. Leaving no one behind: a neglected tropical disease indicator and tracers for the Sustainable Development Goals: Box 1. International Health. 2016 March 3;8(suppl 1):15–8.

19. Essential Package of Health Services: Secondary & Tertiary Care: The District County & National Health Systems, November 2011. Monrovia, Liberia; 2011.

20. Republic of Liberia Investment Plan for Building a Resilient Health System in Liberia. 2015.

21. STRATEGIC PLAN FOR INTEGRATED CASE MANAGEMENT OF NEGLECTED TROPICAL DISEASES (NTDs). 2016.

22. Dean L, Tolhurst R, Nallo G, Kollie K, Bettee A, Theobald S. Neglected tropical disease as a ‘biographical disruption’: Listening to the narratives of affected persons to develop integrated people-centered care in Liberia. PLOS Neglected Tropical Diseases. 2019 September 6;13(9).

23. Republic of Liberia 2008 Population and Housing Census. Monrovia, Liberia; 2008.

24. Collins SE, Clifasefi SL, Stanton J, The LEAP Advisory Board, Straits KJE, Gil-Kashiwabara E, et al. Community-based participatory research (CBPR): Towards equitable involvement of community in psychology research. American Psychologist. 2018 Oct;73(7):884.

25. Zawolo G, Kollie K, Bettee A, Siakeh A, Thomson R, Theobald S, et al. Implementing and integrating NTD programmes in Liberia: Reflections from key stakeholders. Liverpool, UK; 2019 Jan.

26. Kasang C, Krishnan N, Menkor N, Puchner K. Impact of General Health Volunteers on Leprosy Control in Nimba County, Liberia: an experience from the Ganta Leprosy Rehabilitation Centre over 2013-2015. Leprosy Review. 2019;90(2):161–6.

27. Glennie M, Gardner K, Dowden M, Currie BJ. Active case detection methods for crusted scabies and leprosy: A systematic review. PLOS Neglected Tropical Diseases. 2021 July 23;15(7).

28. Buyon L, Slaven R, Emerson PM, King J, Debrah O, Aboe A, et al. Achieving the endgame: Integrated NTD case searches. PLOS Neglected Tropical Diseases. 2018 December 20;12(12).

29. Barogui YT, Sopoh GE, Johnson RC, de Zeeuw J, Dossou AD, Houezo JG, et al. Contribution of the Community Health Volunteers in the Control of Buruli Ulcer in Bénin. PLoS Neglected Tropical Diseases. 2014 October 2;8(10).

30. Vouking MZ, Tamo VC, Mbuagbaw L. The impact of community health workers (CHWs) on Buruli ulcer in sub-Saharan Africa: a systematic review. Pan African Medical Journal. 2013;15.

31. Vouking MZ, Takougang I, Mbam LM, Mbuagbaw L, Tadenfok CN, Tamo CV. The contribution of community health workers to the control of Buruli ulcer in the Ngoantet area, Cameroon. Pan African Medical Journal. 2013;16.

32. Ukwaja KN, Alphonsus C, Eze CC, Lehman L, Ekeke N, Nwafor CC, et al. Investigating barriers and challenges to the integrated management of neglected tropical skin diseases in an endemic setting in Nigeria. PLOS Neglected Tropical Diseases. 2020 April 30;14(4).

33. Succhanda John A, Samuel P, Rao S, Das S. Assessment of needs and quality care issues of women with leprosy. 2010 Mar;81(1):34–40.

34. Ozano K, Dean L, Yoshimura M, MacPherson E, Linou N, Otmani del Barrio M, et al. A call to action for universal health coverage: Why we need to address gender inequities in the neglected tropical diseases community. PLOS Neglected Tropical Diseases. 2020 March 12;14(3).

35. Casulli A. New global targets for NTDs in the WHO roadmap 2021–2030. Plos NTDs. 2021;

